# Evaluating the performance of LYDIA: an AI-powered assistant in detection of metastatic tumors in lymph nodes

**DOI:** 10.1101/2025.05.22.25328136

**Authors:** Georgios Eleftherios Kalykakis, Isaak Tarampoulous, Konstantinos Gyftodimos, Athanasia Sepsa, Giorgos Agrogiannis, Giannis Vamvakaris, Menelaos G Samaras, Christos Spyropoulos, Chrysostomos Manolis, Nikolaos Niotis, Thomas Papathymiopoylos, Carmen van Dooijeweert, Gerben Breimer, Paul J van Diest, Nikolas Stathonikos, Konstantinos Vougas

## Abstract

The integration of AI in histopathology represents a significant advancement in diagnosing metastatic cancers. LYDIA (**LY**mph no**D**e ass**I**st**A**nt) is a commercially available AI-powered tool designed to annotate tumors in lymph node sections to aid histopathologists in diagnosing metastases. This study rigorously evaluates LYDIA’s standalone performance and the time and cost benefits on diagnostic workflow. In this study, LYDIA’s performance was rigorously evaluated on a blind image dataset comprising 366 whole slide images (WSIs) from metastatic breast, colon, lung, and skin cancers. Additionally, a clinical diagnostic workflow was simulated using an internal cohort of 105 WSIs, evaluated by four experienced histopathologists, to assess the impact of LYDIA when used as a decision support tool on diagnostic accuracy and case handling time. A Monte Carlo simulation was also conducted to estimate potential reductions in Immunohistochemistry (IHC) costs associated with AI assistance in sentinel lymph node evaluation. The analysis demonstrated that LYDIA achieved excellent diagnostic accuracy with high ROC-AUC scores (0.995 for breast cancer, 0.963 for colon cancer, 0.973 for lung cancer, and 0.983 for melanoma). In a clinical setting, AI-assisted diagnosis significantly reduced time-to-diagnosis across all metastasis sizes, with a maximum of 1.59-fold acceleration for micro-metastases (saving 26.5 seconds per WSI). LYDIA also enhanced pathologists’ diagnostic performance, increasing their sensitivity from 77.3% to 87.3%. The Monte Carlo simulation indicated an average saving of €7.89 per case, highlighting cost benefits from reduced IHC requests. In conclusion, the findings highlight LYDIA’s capabilities as a supportive tool that holds the potential to reduce diagnostic turnaround times and assist pathologists in identifying important regions within slides. This study illustrates the capability of AI-powered solutions like LYDIA to accelerate diagnosis and ultimately improve patient care. The observed time and cost benefits suggest that integrating AI into routine pathology practice can improve diagnostic quality, reduce costs and optimize resource utilization.

## Introduction

Histopathology plays an instrumental role in diagnosis and therapy of metastatic cancers. Diagnosis of metastases typically entails meticulous examination of lymph node biopsies by trained histopathologists. Traditionally, histopathological examination is performed under a microscope (Roskell & Buley, 2012). However, technological advancements in scanners and computer software facilitated a physical-to-digital transition (Samueli et al., 2024). Currently, diagnosis of metastases primarily relies on the evaluation of whole slide images (WSIs) of H&E-stained lymph node sections on digital platforms by expert pathologists (Jahn et al., 2020). This digital pathology era heralds the opportunity to utilize computational methods in assisting pathologists to detect tumors and expedite the diagnostic process. The high precision and efficiency of AI in extracting information from images has sparked growing interest in applying AI-powered image analysis to support diagnostic workflows in pathology (Dang et al., 2025; Ma et al., 2025). However, despite its promise, deep learning remains in the early stages of clinical adoption—largely due to a lack of trust among pathologists and concerns about being replaced by AI (Nakagawa et al., 2023). Therefore, positioning AI as a decision-support tool rather than a replacement for human expertise is essential. This shift will be driven by rigorous validation in real-world clinical settings.

Recent advances in deep learning, particularly with the introduction of transformer-based architectures and self-supervised learning, have further improved the performance of computational pathology systems on large-scale image data (Lu et al., 2021; Dosovitskiy et al., 2020; Dehaene et al., 2020). Nevertheless, robust, independent, multi-cancer validation studies are still limited, and clinical adoption requires not only high diagnostic accuracy but also demonstrable workflow efficiency and user acceptance (Campanella et al., 2019; Echle et al., 2021). There is a critical need for comprehensive evaluations of commercial AI systems across diverse tumor types and in realistic clinical settings to establish their true utility and limitations in supporting expert diagnosis (Steiner et al., 2018; Baxi et al., 2022).

DeepPath has developed a commercially available AI-powered system named LYDIA (**LY**mph no**D**e ass**I**st**A**nt) (https://www.deeppath.io/deeppath-lydia/). The system, which is commercially available, annotates tumors in lymph node sections to assist histopathologists in diagnosing metastases (a high volume task) faster and with greater accuracy. LYDIA’s unique ability is to point towards the regions that drive its decision and help draw attention to potential macro, micro-metastasis or even isolated tumor cells in lymph node sections.

In this study, we present a comprehensive evaluation of LYDIA, on real-world data. First, we assess its diagnostic performance on a blinded external dataset from the University Medical Center Utrecht (UMCU). Secondly, we simulate a clinical diagnostic workflow using an internal cohort to evaluate the AI’s impact on pathologist accuracy, and diagnostic speed, allowing for a detailed comparison of pathologist performance with and without AI-assistance. Additionally, a cost-benefit analysis was undertaken to quantify the economic implications of integrating AI into routine pathology practice. More broadly, by addressing both LYDIA’s standalone performance on retrospective blinded images and its value as a decision support tool (quantifying the time and cost-benefit of the LYDIA-human interaction) aims to elucidate the practical benefits and limitations of integrating AI decision support in clinical pathology.

## Materials & Methods

### LYDIA System Overview

The LYDIA system operates within a secured facility network and integrates seamlessly with a Picture Archiving and Communication System (PACS). It supports whole slide images (WSI) of H&E structured in a pyramidal format, allowing for analysis at multiple resolutions and zoom levels. The system natively supports a wide range of formats, including .svs, .tif, .dcm, .ndpi, .mrxs, .isyntax, and .bif. To process these images, the system divides the entire slide into a grid, selectively identifies tissue-containing regions while filtering out non-informative areas such as blank background. Detected tissue regions are then passed through a classification pipeline, and the system returns the regions of interest (ROIs) to the user. (Figure 1) Depending on the specific integration, the system can also generate and provide probabilistic heatmaps to enhance interpretability. All processed images and results are managed and stored according to the protocols defined by the PACS.

**Figure 1:**
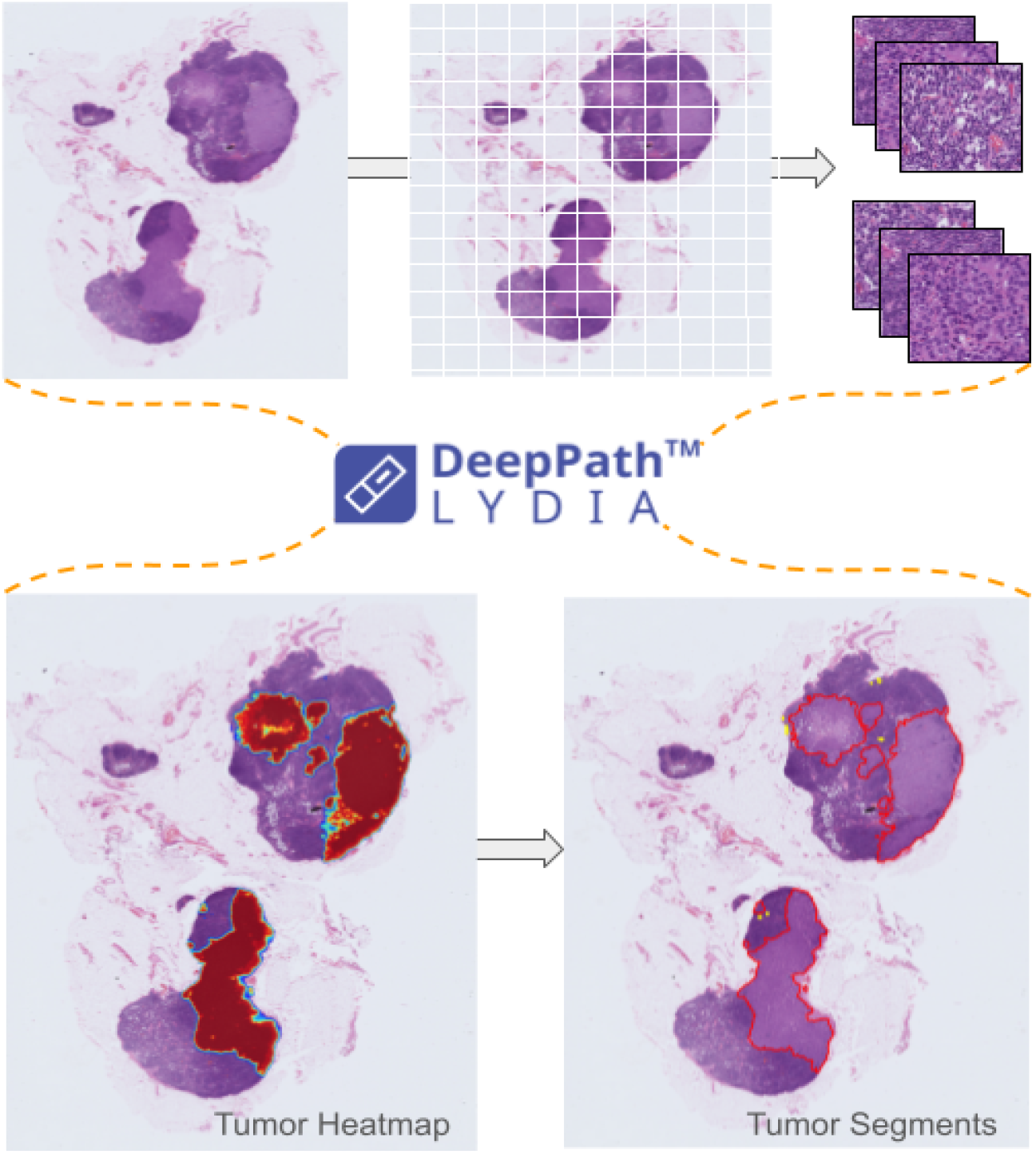
Overview of the LYDIA pipeline. WSIs of H&E-stained lymph nodes are divided into patches and a probability of tumor presence is assigned to each patch. Patch-level predictions are aggregated into tumor heatmaps, which are refined into discrete tumor segments and returned to the user for review.

### Study Cohorts

This study comprises two distinct cohorts: (1) an external single-center cohort used exclusively for performance validation of the system and (2) an internal validation cohort designed to assess time efficiency and diagnostic improvement in clinical practice, with and without AI assistance. The experimental design is outlined in **Figure 2**.

**Figure 2:**
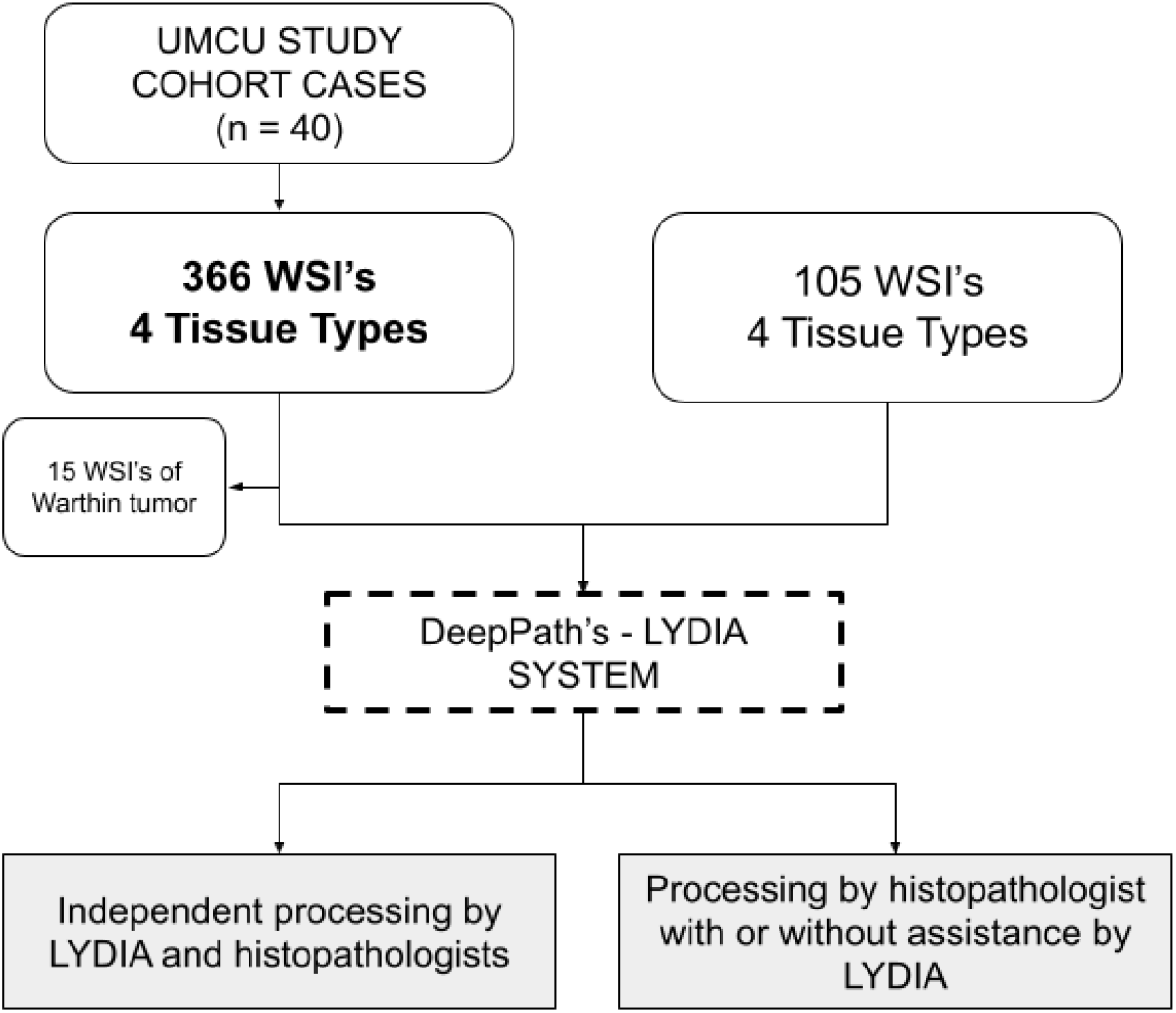
Overview of the two study cohorts used to evaluate LYDIA. The external UMCU cohort (351 WSIs) was used for system performance validation, while the prospective clinical cohort of 105 WSIs assessed diagnostic speed and accuracy with and without AI assistance.

### UMCU study Cohort

In this study cohort, whole slide images of lymph nodes of anonymized cases from metastatic breast, colon, lung, and skin tumors were collected by the University Medical Center Utrecht (UMCU). Cases were gathered from a retrospective cohort based on the initial report which also determined the case-level tumor presence status (Supplementary Material Table S1).

The images were uploaded to an Amazon Web Services (AWS)-based LYDIA deployment (https://umc.deeppath.io/). The processing time and computational resources were recorded for further evaluation. LYDIA generates a probabilistic heatmap of tumor presence and tumor segments are defined by a predefined threshold. (Figure 3).The study was approved by the corresponding local institutional review boards.

**Figure 3:**
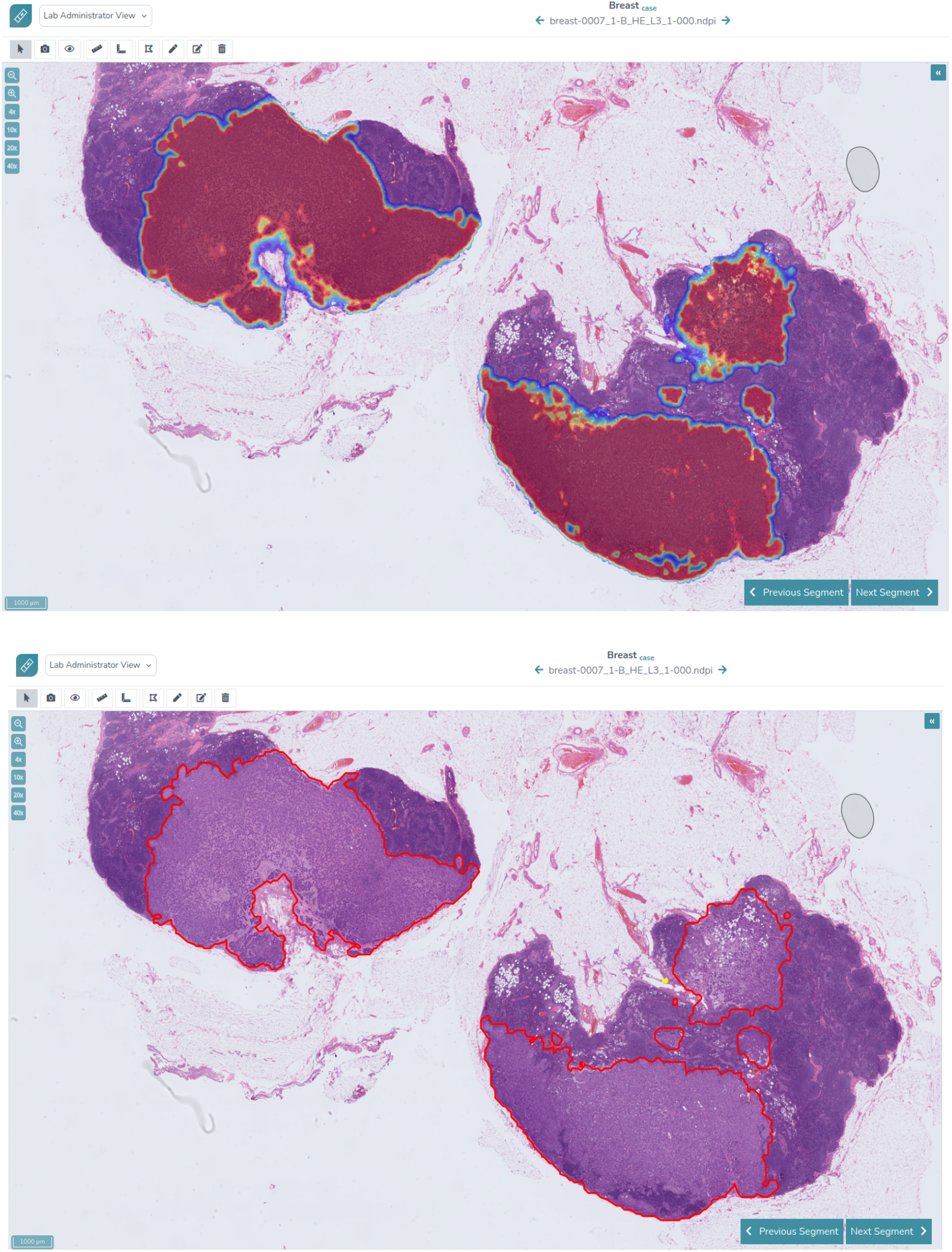
Overview of LYDIA’s user interface. Closed up view displaying (Top) the whole slide image with heatmap annotations and (Bottom) the same whole slide image with Region of interest annotations from LYDIA.

### Prospective Clinical Study Cohort

Four histopathologists, each with more than five years of experience, were tasked with evaluating 105 whole slide images (WSIs) of metastatic tumors from breast, colon, lung, and skin tissues to assess their metastatic status. Positive images where LYDIA demonstrated 100% sensitivity were selected, as the primary objective of the study was to quantify time savings rather than evaluate diagnostic performance. The study was conducted under two distinct scenarios over the course of 4 weeks: one in which the histopathologists performed the task unaided, and another where they utilized the AI-assisted tool, LYDIA. The WSIs were randomly shuffled and rotated between sessions to mitigate memory bias. Diagnostic decisions and time-to-diagnosis were recorded for each scenario and subsequently analyzed for comparison.

### Self-Supervised ViT-Based Feature Extraction and Patch-Level Inference in LYDIA

LYDIA system performs tissue classification analysis by leveraging features extracted from Vision Transformer (ViT) models. Whole slide images (WSIs) pose a significant challenge due to their large size and high dimensionality. To address this complexity, gigapixel compression techniques were employed, enabling efficient processing of these massive images. Specifically, a grid was superimposed onto each gigapixel image, and features were extracted patch-by-patch using an in-house Vision Transformer (ViT). This ViT model was trained using a custom self-supervised framework inspired by the state-of-the-art DINO (Self-Distillation with No Labels, Caron et al., 2021) approach, which facilitates robust unsupervised feature learning, enhanced by aggressive augmentation policies. The result of this process is NeoPath, our in-house lightweight, 22M parameter Foundation Model that powers LYDIA. The generated features are then used as input to downstream machine-learning ensembles, trained under supervised learning to infer the probability of tumor presence. To ensure robust learning, a smart sampling strategy was adopted to balance tumor and healthy tissues across cases, enhancing the model’s ability to generalize across diverse histological morphologies. As such, LYDIA processes the extracted features from individual patches to infer tumor probabilities for each patch. Those probabilities are compiled to form tumor-presence probabilistic heatmaps, out of which the regions of interest (tumor segments) are generated. These ROIs are then overlaid onto the original WSIs, providing pathologists with augmented visualizations that highlight critical areas for inspection.

### Statistical Analysis

Pairwise comparisons were calculated using the Wilcoxon signed-rank test, and a p-value of less than 0.05 was considered statistically significant.

Performance analyses regarding the UMCU cohort study were performed at slide and segment level regarding the DeepPath LYDIA’s performance. Processing time per WSI was calculated in seconds. For slide level metrics, Balanced accuracy (BA), F1 score, True Positive Rate (TPR or Sensitivity), False Positive Rate (FPR), False Negative Rate (FNR) and True negative rate (TNR or Specificity) were calculated comparing the largest annotated tumor area in the ground truth was compared against the maximum predicted patch-level probability within the AI-identified ROI. In negative cases, the highest predicted probability across all patches in the WSI was considered.

Segment level metrics performance was assessed based on the alignment between highlighted potential tumor regions and the actual tumor areas on H&E slides. For Segment level metrics, mean ± Standard Deviation (SD), for Sensitivity and False Positives Per Square Millimeter (FP/mm^2^) were calculated and reported. To put the latest metric into perspective, we internally quantified the average lymph node (LN) area, which was found to be approximately 60 mm2. Regarding the prospective clinical study, median ± 95 CI and percentage time saved per WSI were reported. Balanced accuracy (BA), F1 score,True Positive Rate (TPR), False Positive Rate (FPR), False Negative Rate (FNR) and True negative rate (TNR) were were calculated by comparing cases with the highest metastasis labels to negative cases.

### Monte Carlo Simulation for IHC Cost Reduction Using LYDIA in Sentinel Lymph Node Evaluation

A Monte Carlo simulation was conducted to estimate the potential reduction in Immunohistochemistry (IHC) costs associated with the use of an AI-based decision support system compared to routine diagnostic practice without AI assistance in the evaluation of sentinel lymph node (SLN) slides. The simulation modeled 50000 hypothetical patient cases, each comprising six SLN slides (van Dooijeweert et al., 2024). Slide-level pathological statuses were randomly assigned based on population prevalence rates for breast cancer (van Dooijeweert et al., 2024): Negative (68.9%), Isolated Tumor Cells (ITC, 9.5%), Micro-metastasis (12.6%), and Macro-metastasis (9.0%). Diagnostic performance under each scenario was derived from the confusion matrices generated by the pathologists performance in the time-benefit section of the current study. In the AI-assisted scenario, the probabilities of misclassifying true-positive findings as “Negative” were 1.67% for Macro-metastases, 15% for Micro-metastases, and 30% for ITCs. In the non-AI-assisted scenario, the corresponding misclassification probabilities were 5% for Macro-metastases, 21.67% for Micro-metastases, and 60% for ITCs. In order to compensate for real-world sensitivity of the AI model (99.5% for micro-metastases, 75.5% for ITC) the following correction was applied to the AI-Assisted scenario: micro-correction = (21.67% – 15%) × (100% - 99.5%), itc-correction = (60% – 30%) × (100% - 75.5%). Each slide’s diagnosed status was simulated independently using random sampling weighted by scenario-specific error probabilities.

For determining IHC utilization, two primary protocols were simulated and under both protocols, each IHC-stained slide incurred a fixed cost of €25:

Protocol 1: IHC was requested for all slides within a case if, and only if, all slides in that case were assessed as ‘Negative’ by the respective diagnostic pathway (AI-assisted or non-AI-assisted). If any slide within a case was diagnosed as positive (ITC, Micro-metastasis, or Macro-metastasis), IHC was deemed unnecessary for that case, and the associated IHC cost was set to €0.

Protocol 2: IHC was requested for each slide assessed as ‘Negative’ within a case if, and only if, no slides in that case were diagnosed as containing either Macro-metastasis or Micro-metastasis by the respective diagnostic pathway. If Macro- or Micro-metastasis was detected in any slide of the case, IHC was deemed unnecessary, and the associated IHC cost was €0.

## Results

### Dataset Collection Analytics

We have curated two datasets for the two study cohorts. The UMCU study cohort consists of 351 WSI’s of lymph nodes derived from 39 anonymized cases including metastatic breast, colon, lung, and skin tumors. Patients’ demographics are present in Table 1. Initially, 15 WSI’s from one Melanoma case were excluded because it was a case of Warthin tumor (benign neoplasm). Tissue data distribution and percentage of tumor or benign cases per tissue type are present in Figure 4 A-B.

**Table 1.**
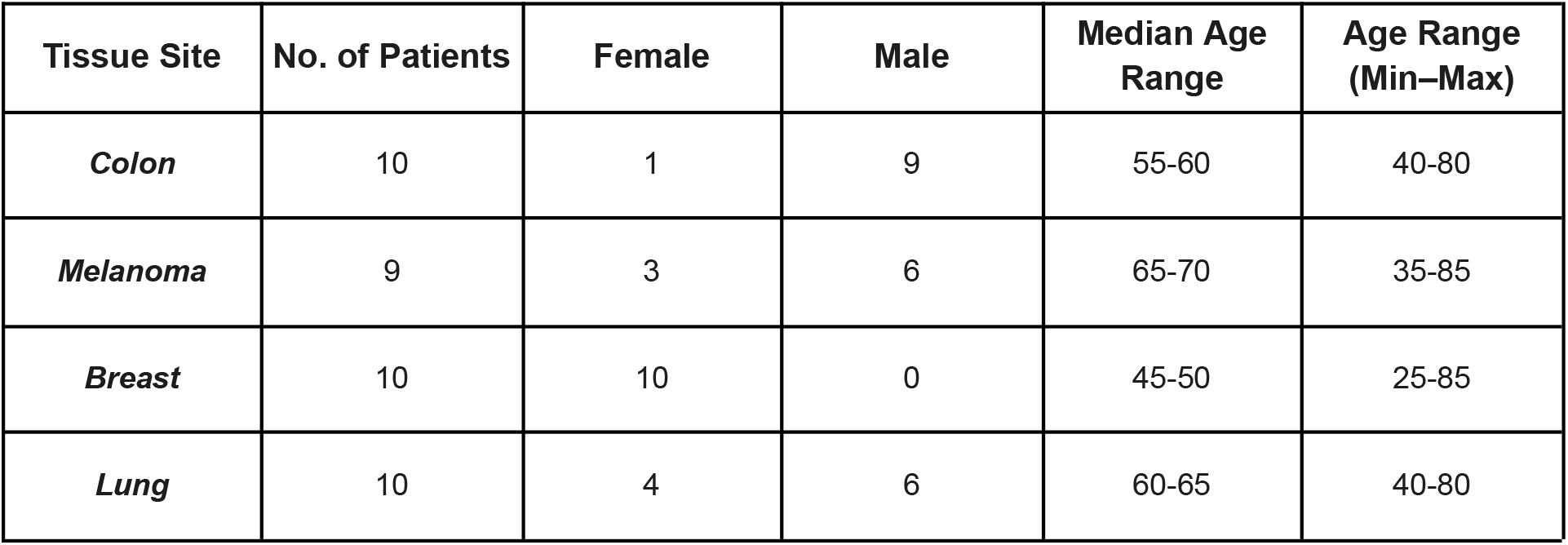
UMCU Patients Demographics

**Figure 4:**
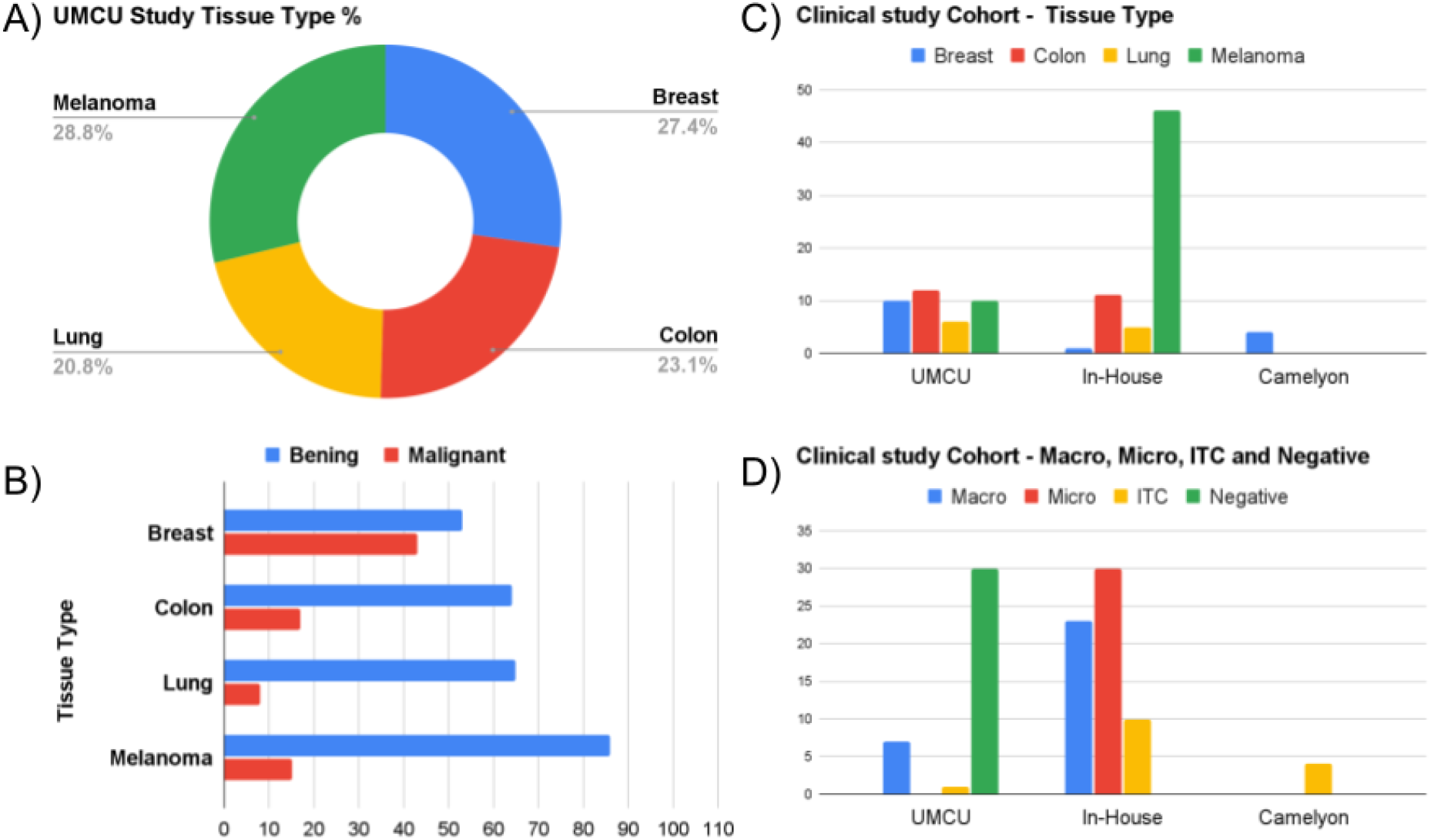
Distribution of Tissue Types and Metastasis Categories Across Study Cohorts. (A) Proportional distribution of tissue types—breast, colon, lung, and melanoma—in the UMCU validation cohort (n = 351 WSIs); (B) Case-level breakdown of benign versus malignant classifications across tissue types in the UMCU cohort; (C) Tissue type distribution across sources in the prospective clinical study cohort, including UMCU, DeepPath in-house, and Camelyon datasets (n = 105 WSIs); (D) Distribution of metastasis grading in the prospective clinical study cohort, categorized as macro-metastasis, micro-metastasis, isolated tumor cells (ITCs), and negative cases.

In addition to that, the Prospective Clinical Study Cohort consists of 105 WSI’s derived from UMCU dataset (38 WSI’s), Camelyon dataset (4 WSI’s) and DeepPath’s in-house dataset (63 WSI’s) covering the same 4 tissue types as UMCU dataset. Details for the tissue distribution and metastasis grading are present in Figure 4 C-D.

### Evaluation of LYDIA’s performance in detecting metastatic tumors

We first evaluated the model’s performance in detecting metastatic tumors from breast, colon, lung tumors and melanoma in WSIs from lymph nodes regardless of the tumor of origin utilizing the UMCU study cohort.

Regarding whole slide images level metrics (WSI) at the threshold calibrated for 99% sensitivity,LYDIA’s whole slide image (WSI) performance metrics demonstrated strong diagnostic capability across all four cancer types. The ROC-AUC was highest for breast cancer (0.995), followed by melanoma (0.983), lung (0.973), and colon (0.963). Balanced accuracy (BA) at 99% sensitivity was 0.915 for breast, 0.938 for colon, 0.923 for lung, and 0.925 for melanoma. F1 scores were highest for breast cancer (0.905), followed by colon (0.800), melanoma (0.711), and lung (0.615). False positive rates (FPR) at this sensitivity threshold were lowest for melanoma (0.149) and highest for breast (0.170), with colon and lung at 0.123 and 0.154, respectively. True negative rates (TNR, specificity) were 0.830 for breast, 0.877 for colon, 0.846 for lung, and 0.851 for melanoma (Table 2).

**Table 2.** LYDIA WSI-level performance characteristics at >99% sensitivity.

|  | **Breast** | **Colon** | **Lung** | **Melanoma** |
| --- | --- | --- | --- | --- |
| *ROC-AUC* | 0.995 | 0.963 | 0.973 | 0.983 |
| *Balanced accuracy (BA)* | 0.915 | 0.938 | 0.923 | 0.925 |
| *F1 score* | 0.905 | 0.8 | 0.615 | 0.711 |
| *False positive rate (FPR)* | 0.17 | 0.123 | 0.154 | 0.149 |
| *True negative rate (TNR, Specificity)* | 0.83 | 0.877 | 0.846 | 0.851 |

**Table 3.**
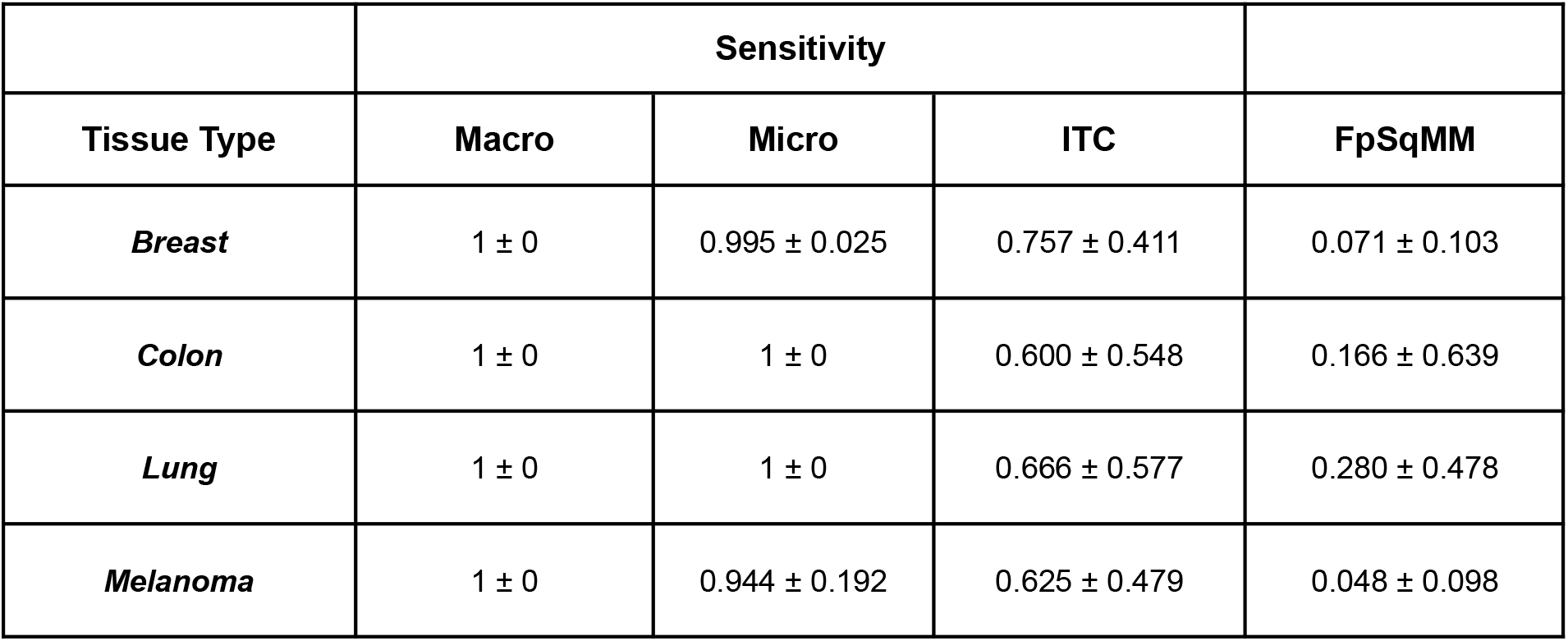
LYDIA Segment Level performance characteristics.

|  | **Sensitivity** |  |  |  |
| --- | --- | --- | --- | --- |
| **Tissue Type** | **Macro** | **Micro** | **ITC** | **FpSqMM** |
| ***Breast*** | 1 ± 0 | 0.995 ± 0.025 | 0.757 ± 0.411 | 0.071 ± 0.103 |
| ***Colon*** | 1 ± 0 | 1 ± 0 | 0.600 ± 0.548 | 0.166 ± 0.639 |
| ***Lung*** | 1 ± 0 | 1 ± 0 | 0.666 ± 0.577 | 0.280 ± 0.478 |
| ***Melanoma*** | 1 ± 0 | 0.944 ± 0.192 | 0.625 ± 0.479 | 0.048 ± 0.098 |

LYDIA demonstrated robust segment-level performance across metastasis sizes and tissue types. For breast cancer, the model achieved perfect sensitivity (1.00 ± 0.00) for macro-metastases, maintained high sensitivity for micro-metastases (0.995 ± 0.025), and showed moderate detection of isolated tumor cells (ITCs) (0.757 ± 0.411), while maintaining low false positives per square millimeter (FpSqMM) values (0.071 ± 0.103). In colon cancer, sensitivity was perfect for macro- and micro-metastases (1.00 ± 0.00) but substantially lower for ITCs (0.600 ± 0.548), accompanied by slightly elevated FpSqMM (0.166 ± 0.639). Lung cancer segments exhibited perfect sensitivity for macro- and micro-metastases (1.00 ± 0.00) with acceptable ITC detection (0.667 ± 0.577), with the highest FpSqMM observed in this group (0.280 ± 0.478). Melanoma segments showed near-perfect sensitivity for macro-metastases (1.00 ± 0.00) and strong detection of micro-metastases (0.944 ± 0.192), with moderate ITC identification (0.625 ± 0.479), while maintaining the lowest FpSqMM values (0.048 ± 0.098) among all tissue types (Table 4).

**Table 4.** Pathologists’ diagnostic performance with and without LYDIA assistance in prospective clinical study.

|  | **All Sizes** |  | **Macro** |  | **Micro** |  | **ITC** |  |
| --- | --- | --- | --- | --- | --- | --- | --- | --- |
|  | **Pathologists** | **Pathologists + LYDIA** | **Pathologists** | **Pathologists + LYDIA** | **Pathologists** | **Pathologists + LYDIA** | **Pathologists** | **Pathologists + LYDIA** |
| ***Balanced accuracy (BA)*** | 83.25% | 89.91% | 92.08% | 95.41% | 83.75% | 88.75% | 64.58% | 81.25% |
| ***F1 score*** | 85.13% | 91.76% | 92.30% | 95.54% | 82.81% | 88.31% | 49.48% | 75.67% |
| ***True positive rate (TPR, Sensitivity)*** | 77.33% | 87.33% | 95.00% | 98.33% | 78.33% | 85.00% | 40.00% | 70.00% |
| ***False positive rate (FPR)*** | 10.83% | 7.50% | 10.83% | 7.50% | 10.83% | 7.50% | 10.83% | 7.50% |
| ***False negative rate (FNR)*** | 22.67% | 12.67% | 5.00% | 1.67% | 21.67% | 15.00% | 60.00% | 30.00% |
| ***True negative rate (TNR, Specificity)*** | 89.17% | 92.50% | 89.17% | 92.50% | 89.17% | 92.50% | 89.17% | 92.50% |

### Characterization of LYDIA processing time and resources

Whole-slide images (WSIs) of H&E-stained lymph nodes were processed using Amazon Web Services (AWS) EC2 G4 instances equipped with NVIDIA T4 GPUs and custom Intel Cascade Lake CPUs (https://aws.amazon.com/ec2/instance-types/g4/). The average processing time per WSI, regardless of tumor origin, was approximately 3 minutes (180.73 ± 92.34 seconds). A more granular analysis revealed consistent processing durations across different tumor origins (Figure 5, Table S2). WSIs from breast cancer cases required an average of 208.06 ± 68.73 seconds, with positive and negative cases showing comparable durations (203.83 ± 63.14 seconds and 217.81 ± 80.54 seconds, respectively). Colon cancer WSIs had an average processing time of 192.05 ± 123.28 seconds, with slightly higher durations in positive cases (205.53 ± 132.05 seconds) compared to negative ones (144.85 ± 69.94 seconds). Lung cancer WSIs were processed more rapidly overall, averaging 125.09 ± 91.75 seconds, with positive (128.83 ± 119.57 seconds) and negative (123.59 ± 79.19 seconds) cases demonstrating similar timing. WSIs from melanoma cases required an average of 185.66 ± 65.16 seconds, again with minimal variation between positive (188.56 ± 65.54 seconds) and negative (184.23 ± 65.39 seconds) cases. Importantly, LYDIA supports parallelized WSI processing, enabling the system to allocate slides across multiple GPUs and thereby significantly reduce the per-case runtime. This architecture ensures scalability and responsiveness in high-throughput diagnostic settings.

**Figure 5:**
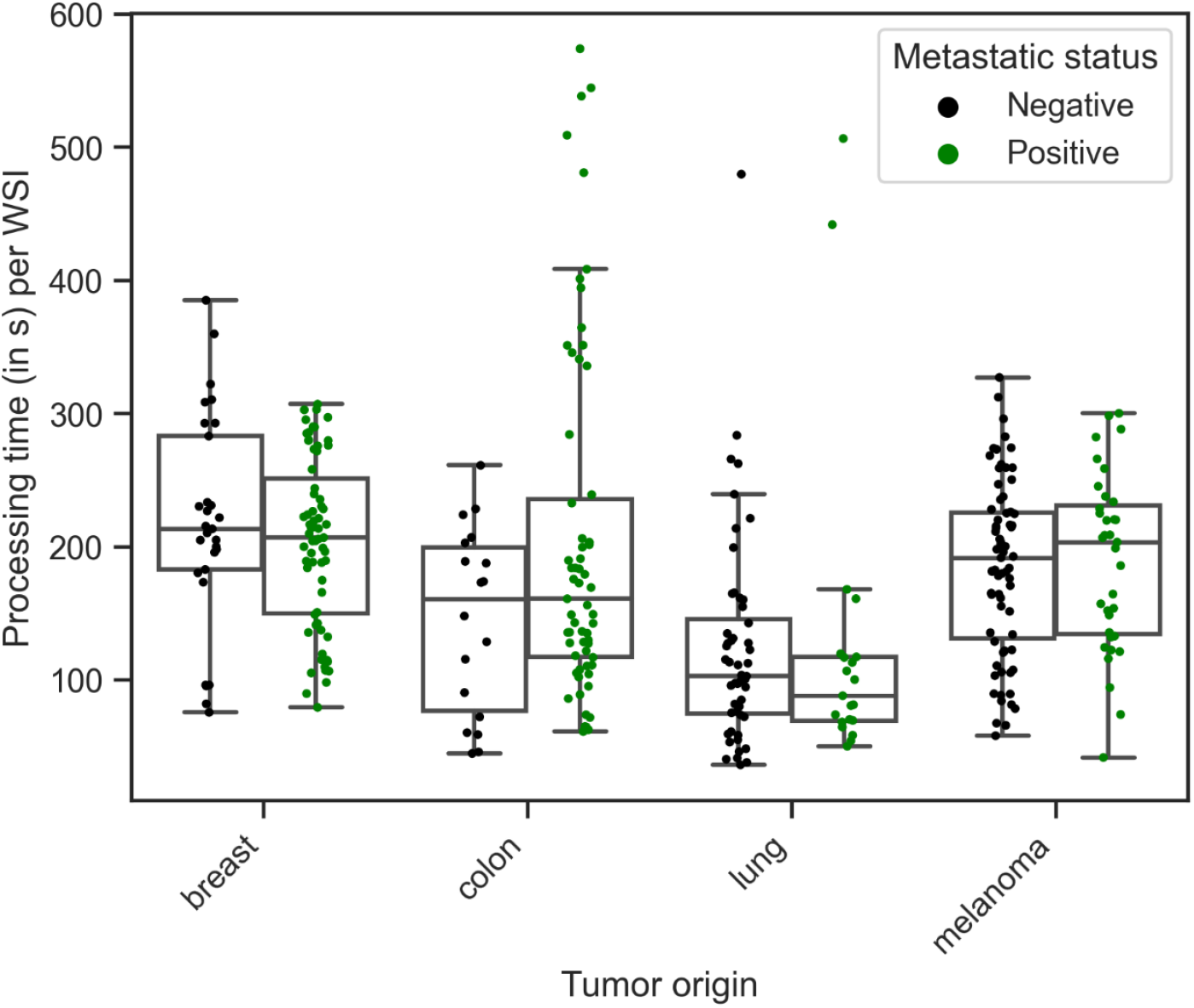
LYDIA’s WSI Processing Times by Tissue Type and Tumor Status. Average processing times (in seconds) for H&E-stained lymph node WSIs across breast, colon, lung, and melanoma cases, stratified by tumor status (positive vs. negative). Analysis was performed on AWS EC2 G4 instances equipped with NVIDIA T4 GPUs. Minimal variation was observed between positive and negative cases within each tissue type, and lung WSIs exhibited the shortest average processing times. The system’s architecture supports parallelized slide allocation across GPUs, enabling scalable performance for high-throughput diagnostic applications.

### Benefit when system used as a decision support tool to diagnostic time and expert performance

Next, we investigated the time benefit and diagnostic performance of LYDIA in a clinical-like environment. To this end, 100 whole slide images were independently evaluated by four experienced histopathologists over a four-week period. For macro-metastases, AI-assisted diagnosis time was significantly reduced compared to non-AI assistance, achieving a 1.3-fold acceleration (saving 4 seconds (−30.7-63.1 seconds p < 0.001). This time-saving effect was consistently observed for micro-metastases (1.59-fold faster; saving 26.5 seconds (−49.1-150.6 seconds) per WSI, p < 0.001), isolated tumor cells (1.27-fold faster; saving 14.5 seconds (−40.2-159.5), p < 0.001), and negative cases (1.41-fold faster; saving 17 seconds (−34.1-165.1 seconds, p < 0.001, Figure 6A).

**Figure 6:**
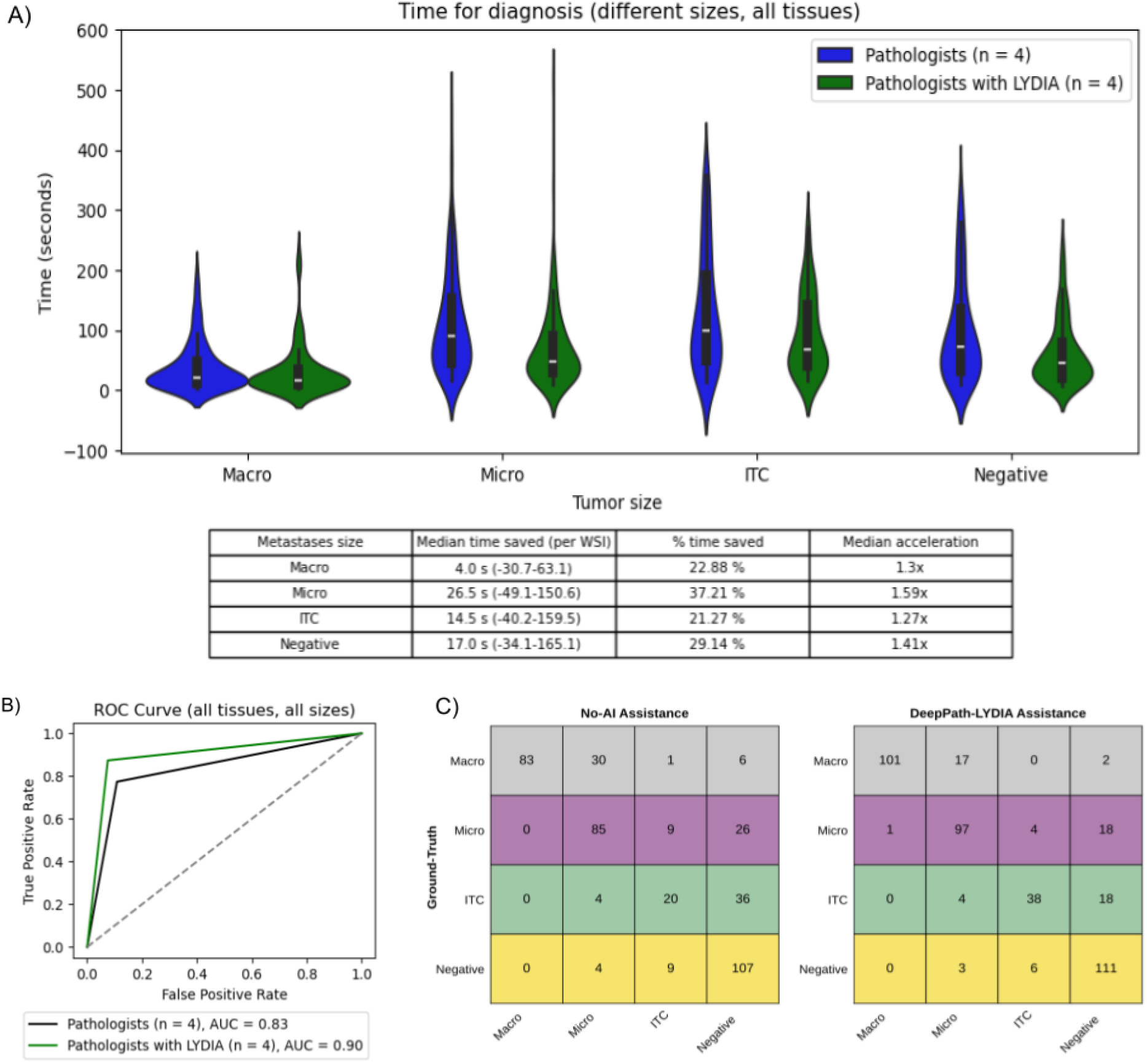
Impact of LYDIA on Diagnostic Time and Accuracy. (A) Time-to-diagnosis per WSI for macro-metastases, micro-metastases, isolated tumor cells (ITCs), and negative cases, comparing AI-assisted and unassisted pathologist performance. Across all categories, AI assistance significantly reduced diagnostic time, with the largest time savings observed for micro-metastases; (B) Improvement in diagnostic accuracy withLYDIA support. Receiver operating characteristic (ROC) AUC increased from 83% to 90%; (C) Confusion matrices indicating pathologists diagnostic accuracy with and without AI assistance for Macro, Micro, ITC and Negative cases.

In addition to accelerating diagnosis, LYDIA significantly enhanced pathologists’ diagnostic performance, increasing the area under the receiver operating characteristic curve (AUC) from 83% to 90% when assisted by the system (Figure 6B). Detailed analysis revealed that for macro-metastases, balanced accuracy (BA) improved from 92.08% to 95.42%, accompanied by an increase in the F1 score from 92.3% to 95.5%. The true positive rate (TPR) rose from 95.00% to 98.33%, while the false positive rate (FPR) decreased from 10.83% to 7.50%. For micro-metastases, BA increased from 83.75% to 88.75%, the F1 score improved from 82.8% to 88.3%, TPR increased from 78.33% to 85.00%, and the false negative rate (FNR) decreased from 21.67% to 15.00%. The most pronounced improvement was observed in ITC detection, where BA increased markedly from 64.58% to 81.25%, the F1 score rose substantially from 49.5% to 75.7%, TPR improved from 40.00% to 70.00%, and FNR was halved from 60.00% to 30.00% (Table 4).

### Cost savings from reduced IHC requests from Monte Carlo Simulation

The Monte Carlo simulation (50000 repeats) evaluated the economic impact of AI assistance in breast cancer SLN assessment under two IHC request protocols. Under Protocol 1 (IHC if all slides per case were assessed as Negative), the AI-assisted pathway averaged an IHC cost of €25.38 per case, compared to €33.27 for the non-AI pathway. This yielded an absolute saving of €7.89 per case (23.72 % relative saving) utilized AI-assistance. Under Protocol 2 (IHC for Negative-assessed slides if no Macro- or Micro-metastasis was detected per case), the AI-assisted pathway averaged €37.71 per case, versus €42.22 for the non-AI pathway. This resulted in an average absolute saving of €4.51 per case (10.69 % relative saving) with AI (Figure 7).

**Figure 7:**
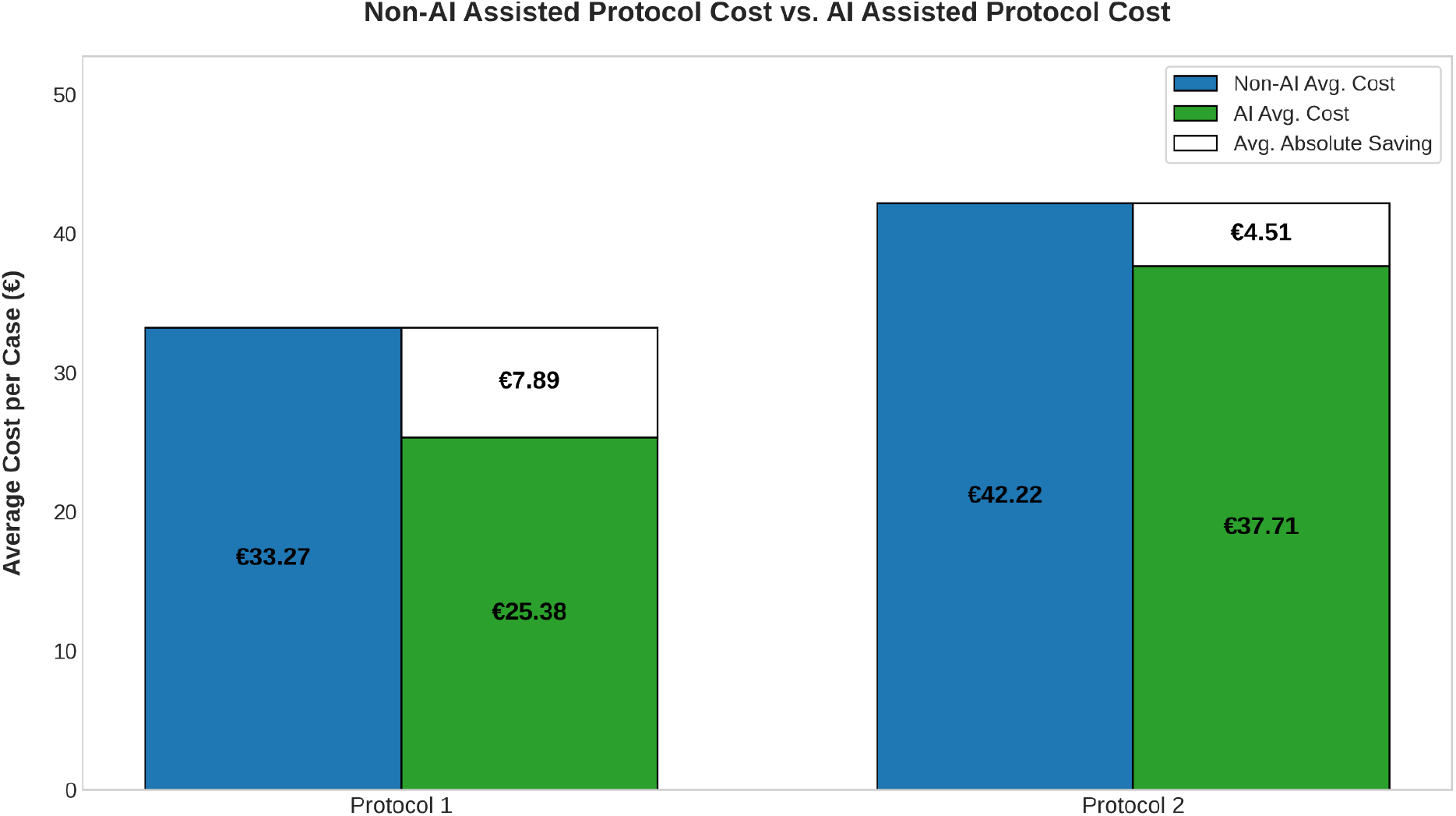
Cost Savings from AI-Assisted Pathway in Breast Cancer Assessment. Comparison of IHC Request Protocols utilizing Monte-Carlo Simulations

## Discussion

Digitization of histopathological slides by the introduction of whole slide scanners in the clinical settings and digital evaluation of them has led to significant time savings (Hanna et al., 2019). The emergence of artificial intelligence promises to further accelerate diagnosis while maintaining quality and reducing costs (Jeong et al., 2025).

This study provides a comprehensive evaluation of LYDIA, an AI-powered tool for the detection of metastatic tumors in lymph node WSIs, in 4 tissue types: breast, lung, colon and skin, across two independent scenarios: a blinded external validation using the UMCU dataset and a prospective clinical study assessing real-world utility for decision support. The LYDIA system performed strongly on the external UMCU dataset across all tested cancer types. It consistently detected macro- and micro-metastases with high sensitivity. Detection of isolated tumor cells was more variable, reflecting the inherent difficulty of these subtle findings. False positive rates differed by tissue type, likely due to morphological complexity. One notable case involved a benign Warthin tumor misclassified as malignant, highlighting a need for refinement in distinguishing neoplastic but non-metastatic tissue. In the prospective clinical study, AI assistance significantly enhanced pathologist performance. Diagnostic accuracy and consistency improved across key metrics. The system especially boosted detection of micro-metastases and isolated tumor cells. Turnaround times were notably reduced with AI support. These findings highlight AI’s value in speeding up and strengthening diagnostic workflows.

These findings are aligned with prior studies that have shown AI can reduce diagnostic turnaround times and improve consistency in challenging diagnostic scenarios, such as the identification of isolated tumor cells (Steiner et al., 2018). The system’s utilization of advanced techniques, such as a self-supervised Vision Transformer (ViT)-based foundation model (NeoPath), likely contributes to this strong performance, aligning with recent advancements in the field field (Dosovitskiy et al., 2020; Chen et al., 2023).

The observed time savings in our study are comparable to those reported by van Dooijeweert et al. (2024). That same study also estimated cost savings of €15 per breast cancer case due to reduced IHC requests—approximately double the maximum value of €7.98 that we calculated using our simulation method. This discrepancy is likely attributable to differences in baseline diagnostic performance: van Dooijeweert et al. reported a pathologist sensitivity of 50%, increasing to 60% with AI assistance, whereas in our study, sensitivity improved from 77% to 87%.

All the above results suggest that integrating AI into routine pathology practice has the potential not only to improve diagnostic quality but also to optimize resource utilization-a key consideration for healthcare systems facing increasing demands and limited specialist availability (Baxi et al., 2022).

LYDIA supports a scalable deployment in the cloud and can analyze every WSI in about 3 minutes. Independent processing of each WSI by a GPU allows for parallel processing of multiple WSIs in batches. This promises to accelerate the overall processing time by increase of batch size, which is only limited by the number of available GPUs. In this study, we demonstrate that LYDIA’s model achieves a remarkable area under the receiver-operating curve (ROC-AUC) across all tested cancer types. The ability of a single model to detect multiple types of metastatic tumors demonstrates LYDIA’s universality, which may potentially be expanded to all types of metastatic tumors.

Testing of LYDIA in a clinical-like environment demonstrated its ability to accelerate diagnosis, in alignment with previous findings from AI-augmented histopathology systems (Baidoshvili et al., 2023). This independent evaluation of LYDIA exemplifies the potential of AI to augment the traditional practices of histopathology. The reliable detection of metastatic tumors by LYDIA may accelerate diagnosis by histopathologists, while increasing precision. However, follow-up studies and cost benefit analysis are needed to quantify the advantage of LYDIA and similar solutions in a clinical setting.

## Limitations and Future Directions

We would like to note this study was limited to a single commercial AI system and, for the clinical workflow component, to a single institution and a select group of pathologists. These factors may limit the generalizability of the findings, underscoring the need for future prospective, multi-center studies and benchmarking across diverse AI solutions. Additionally, the retrospective nature of the validation cohort and the use of simulated cost analysis, while informative, should be complemented by prospective health economic studies in real-world clinical practice.

## Conclusion

The integration of artificial intelligence (AI) into the field of histopathology, particularly through tools like LYDIA, marks a transformative shift in the diagnosis and management of metastatic cancers. This study’s evaluation of LYDIA’s performance in analyzing whole slide images (WSIs) of lymph nodes from patients with various types of cancer has demonstrated its high efficiency and precision. The robust diagnostic performance and workflow benefits observed in this study suggest that future guidelines could formally recommend the use of validated AI-powered decision support systems, like LYDIA, as adjuncts to routine histopathological assessment of lymph nodes in cancer patients. By reducing overall diagnostic turnaround time and unnecessary IHC staining (leading to direct cost savings) without compromising diagnostic accuracy, AI-assisted workflows could be incorporated into guidelines to optimize resource utilization and focus IHC on diagnostically ambiguous cases.

## Supporting information

Supplementary Matterial

## Data Availability

The image dataset used to evaluate the performance of DeepPATH(TM)-LYDIA originates from anonymized patient cases, but it can be made available upon reasonable request for review purposes.

