## Supplementary Matterial for "Evaluating the performance of LYDIA: an AI-powered assistant in detection of metastatic tumors in lymph nodes"

### Supplementary material

#### Supplementary Figures

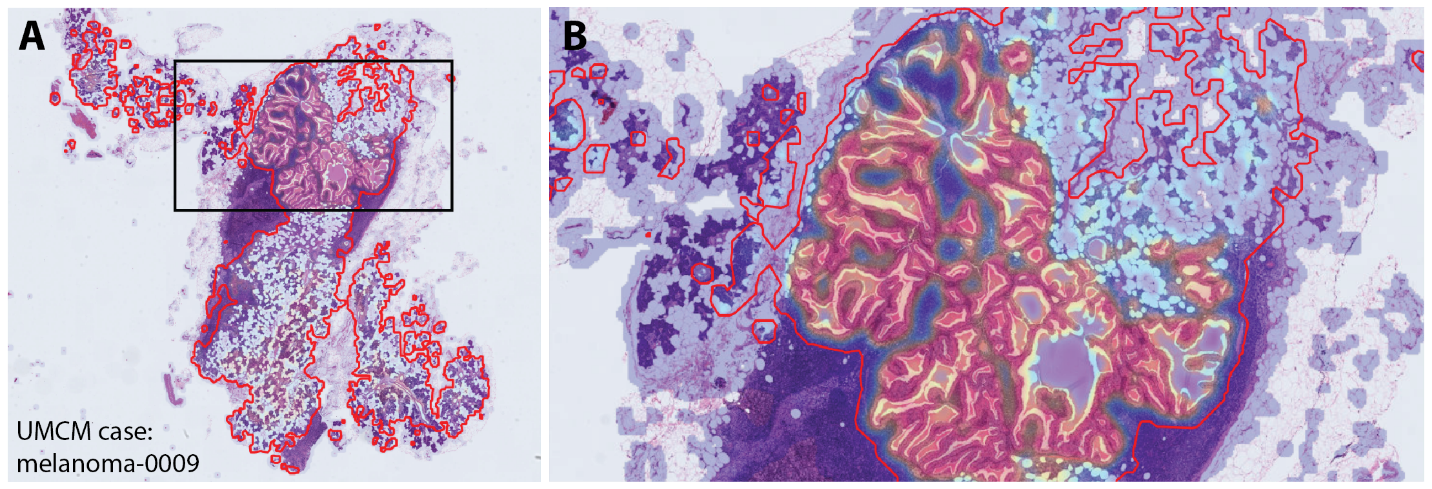

**Figure S1:** (A) Image panel displaying LYDIA’s output for the melanoma-0009 case from the UMCU, demonstrating LYDIA’s ability to detect Warthin tumors (benign neoplasms); (B) Zoomed-in view of the region in which the neoplasm was detected, marked by the black box in (A).

#### Supplementary Tables

**Table S1:** Breakdown of metastasis subtype and WSI count per case in the UMCU validation cohort

| **Case** | **Macro** | **Micro** | **ITC** | **Negative** | **Total** |
| --- | --- | --- | --- | --- | --- |
| *breast-0001* | 0 | 0 | 0 | 5 | 5 |
| *breast-0002* | 10 | 0 | 0 | 5 | 15 |
| *breast-0003* | 0 | 0 | 0 | 5 | 5 |
| *breast-0004* | 0 | 0 | 0 | 5 | 5 |
| *breast-0005* | 0 | 0 | 0 | 5 | 5 |
| *breast-0006* | 0 | 0 | 0 | 6 | 6 |
| *breast-0007* | 5 | 0 | 0 | 5 | 10 |
| *breast-0008* | 16 | 3 | 0 | 11 | 30 |
| *breast-0009* | 0 | 1 | 3 | 1 | 5 |
| *breast-0010* | 4 | 1 | 0 | 5 | 10 |
| *colon-0001* | 0 | 1 | 1 | 8 | 10 |
| *colon-0002* | 3 | 0 | 1 | 9 | 13 |
| *colon-0003* | 0 | 0 | 0 | 1 | 1 |
| *colon-0004* | 0 | 0 | 0 | 6 | 6 |
| *colon-0005* | 8 | 0 | 0 | 19 | 27 |
| *colon-0006* | 1 | 0 | 0 | 5 | 6 |
| *colon-0007* | 0 | 0 | 0 | 3 | 3 |
| *colon-0008* | 1 | 1 | 0 | 5 | 7 |
| *colon-0009* | 0 | 0 | 0 | 7 | 7 |
| *colon-0010* | 0 | 0 | 0 | 1 | 1 |
| *lung-0001* | 2 | 0 | 0 | 5 | 7 |
| *lung-0002* | 0 | 0 | 0 | 10 | 10 |
| *lung-0003* | 0 | 0 | 0 | 6 | 6 |
| *lung-0004* | 2 | 2 | 0 | 8 | 12 |
| *lung-0005* | 0 | 0 | 0 | 14 | 14 |
| *lung-0006* | 2 | 0 | 0 | 0 | 2 |
| *lung-0007* | 0 | 0 | 0 | 3 | 3 |
| *lung-0008* | 0 | 0 | 0 | 6 | 6 |
| *lung-0009* | 0 | 0 | 0 | 5 | 5 |
| *lung-0010* | 0 | 0 | 0 | 8 | 8 |
| *melanoma-0001* | 0 | 0 | 0 | 12 | 12 |
| *melanoma-0002* | 0 | 0 | 0 | 12 | 12 |
| *melanoma-0003* | 0 | 0 | 0 | 6 | 6 |
| *melanoma-0004* | 0 | 0 | 0 | 6 | 6 |
| *melanoma-0005* | 0 | 0 | 0 | 6 | 6 |
| *melanoma-0006* | 2 | 0 | 0 | 6 | 6 |
| *melanoma-0007* | 0 | 0 | 0 | 24 | 24 |
| *melanoma-0008* | 1 | 0 | 0 | 7 | 8 |
| *melanoma-0010* | 0 | 12 | 0 | 7 | 19 |

### **Table S2:** Mean processing times per WSIs for metastatic breast, colon, lung tumors and melanoma.

| **Tumor origin** | **Metastatic Status** | **Processing time** |
| --- | --- | --- |
| Breast | Total | 208.06 ± 68.73 seconds |
|  | Positive only | 203.83 ± 63.14 seconds |
|  | Negative only | 217.81 ± 80.54 seconds |
| Colon | Total | 192.05 ± 123.28 seconds |
|  | Positive only | 205.53 ± 132.05 seconds |
|  | Negative only | 144.85 ± 69.94 seconds |
| Lung | Total | 125.09 ± 91.75 seconds |
|  | Positive only | 128.83 ± 119.57 seconds |
|  | Negative only | 123.59 ± 79.19 seconds |
| Melanoma | Total | 185.66 ± 65.16 seconds |
|  | Positive only | 188.56 ± 65.54 seconds |
|  | Negative only | 184.23 ± 65.39 seconds |
